# Quality of Drug Prescription and Dispensing Practices in a Teaching Hospital in a developing country

**DOI:** 10.1101/2023.03.03.23286781

**Authors:** Owhondah King Samuel, Zite Zorte, Ogaji Daprim Samuel

## Abstract

**Background:** The World Health Organization recommends rational drug usage to protect patient health and quality of life. Study assessed the quality of drug prescription and dispensing in a tertiary hospital.

**Methods:** Cross-sectional study with retrospective and prospective data collection underpinned by the WHO core prescribing indicators. A cluster sample of 10 clinical units and their attached pharmacies in the hospital. Six hundred prescriptions from the pharmacy over six months were randomly selected to evaluate the prescription indicators, 330 patient encounters observed for patient care indicators, and 48 randomly selected doctors to evaluate factors influencing their prescribing practices across the 10 clinics. Descriptive analysis was performed using the Statistical Package for the Social Sciences (SPSS) version 25 and comparison done across all indicators of rational drug use.

**Results:** Average drugs per encounter was 3.4 ± 1.9 drugs. Antibiotics and injections were prescribed in 40.2% and 24.8% encounters respectively. Generic names were used in 43.6% and 97.1% of prescription were from the essential medicine list. Average time for consultation was 17.5 ± 8.0 minutes, dispensing time was 7.7 ± 3.8 minutes, 99.8% of medications were properly labelled, and 82% of patients understood the drug doses. The pharmacies stocked 93.3% of key drugs but no consultation room had the essential drug list. Only 43.7% of physicians demonstrated accurate understanding of rational drug use.

**Conclusion:** Findings on the WHO core drug indicators showed poor quality of drug prescription. Brand prescription, polypharmacy, and antibiotic overuse observed requires intensifying training and monitoring on rational drug use.

## Introduction

Drugs are part of the main therapeutic interventions used in healthcare, for patient care. Drug therapy (pharmacotherapy) is important and relies on the science of pharmacology for continual advancement and on pharmacy for appropriate management.^1^ The majority of patients’ trips to clinics and exposure to healthcare surroundings result in an appropriate drug prescription. A drug prescription is a formal written order from a prescriber to a dispenser in most circumstances, especially in established hospital settings, after complete consultation, examination, and diagnosis. It is a medico-legal document that should be written legibly, accurately, and with completeness.^2^

Rational drug use (RDU) is defined by the World Health Organization (WHO) as “correct prescribing, appropriate distribution, and appropriate patient use of drugs for illness diagnosis, prevention, mitigation, and treatment.”^3^ The patient will receive medicine that is appropriate for his clinical condition, at the correct dose, and at the right time, while also considering the drug’s total cost. According to the World Health Organization (WHO), up to 50% of pharmaceuticals are prescribed, supplied, or marketed inappropriately over the world^3^. Irrational drug usage not only wastes money spent on this category of goods, but also poses a considerable risk to the patient, including the likelihood of drug therapy side effects.^1-4^

The World Health Organization (WHO), in collaboration with the International Network for the Rational Usage of Drugs (INRUD), developed a set of standardized core prescribing and patient care indicators to assess drug use trends in health-care facilities’ outpatient settings.^3^ Each core indicator is made up of five parts. Among the prescription indicators are the degree of polypharmacy, the proportion of pharmaceuticals prescribed with a generic name, the percentage of contacts with at least one antibiotic and injectable, and the percentage of medications indicated from the EDL. Patient care indicators also include the average consultation time, average dispensing time, percentage of medications delivered and labeled, and percentage of patients who know how to take the correct dosage. The WHO recommends a value of 2 (1.4-1.8) for average number of pharmaceuticals per encounter, 30% (20-26.8%) for percent encounters with antibiotics, and 25% (13.4-24.1%) for percent encounters with injection(s), whereas the best value for prescription by generic name and from EDL is 100%. The average consultation time (>10 minutes), average dispensing time (>180 seconds), and the proportion of medications given, labeled, and patient knowledge should all be at least 100% according to WHO patient care indicators.^3-7^

Several authors have written about the issue of irrational drug usage in Africa and around the world. In Pakistan, doctors only spent 1.2 minutes with patients in consultation rooms, there was increased brand prescription, and polypharmacy and antibiotic over-prescribing were prevalent problems.^8^ In Ethiopia, drug prescriptions were not correctly labeled for patients, and prescribers overprescribed antibiotics. Dispensers spent only 59 seconds with patients, resulting in inadequate drug education.^9^ In Nepal, over 37% of antibiotics prescribed in 2017 were unnecessary, leading in additional money being squandered.^10^ This irrationality in drug use has an impact not just on patients but also on entire countries. According to reports, 40% of Nepalese medical spending was squandered in 2017 owing to incorrect prescriptions.^10^

Polypharmacy, poor generic prescribing, and antibiotic usage have all been highlighted as important issues in Nigeria, according to studies conducted in two Nigerian teaching hospitals in Lagos.^11^ Another investigation in Nigeria in 2003 looked at drug use and antibiotic prescriptions and showed that, contrary to WHO recommendations, the ratio of antibiotics per prescription in public hospitals was above 75% and 55 percent in private hospitals.^12^ Antibiotic resistance, higher treatment costs, and further patient impoverishment will result from such bad prescribing patterns, putting the family’s and community’s financial well-being at risk. More than half of patients do not take drugs appropriately even when they are available, according to the WHO’s drug perspectives, resulting in harmful side effects.^3^ Incorrect self-medication, limited efficacy (e.g., under-therapeutic dosage of tuberculosis or leprosy drugs), antibiotic resistance (due to widespread overuse and under-therapeutic dosage of antibiotics), drug dependence (due to daily use of painkillers and tranquilizers), and infection risk are all medical outcomes of such irrational use (due to improper injection use). Injections are responsible for abscesses, hepatitis B, and HIV/AIDS.^3,7^

Overprescribing, multi-drug prescribing, drug misuse, unnecessary expensive medicine use, and overuse of antibiotics and injections are among the most common drug-use concerns, according to studies.^3,13,14^ This was the rationale for the conduct of this research aimed at determining the state of rational drug use in the University of Port Harcourt Teaching Hospital.

Adoption of the WHO standard on drug prescription and patient care serves as a quality control measure that can be used to improve drug prescription and dispensing in tertiary health facilities. This study ascertained the pattern of drug prescription, evaluate patient care in relation to contact time and drug information, evaluate the availability of essential drugs and identify the factors that influence drug prescribing in the University of Port Harcourt Teaching Hospital.

### Methodology

This study was conducted in the University of Port Harcourt Teaching Hospital (UPTH) which is a tertiary hospital in Port Harcourt, Rivers State. This study adopted a descriptive cross-sectional study design. Prescribing encounters dispensed within six months (June 1, 2021-Nov 30, 2021) served as the data source among which 600 prescriptions were drawn for the actual study using a sampling fraction of 1:3. For the patient care indicator, the patients’ folder served as the data source, while the doctors in the consulting clinics served as the population for studying factors associated with prescription. The sample size for this study was calculated using the Cochran’s formula for minimum sample size of 323 was determined using the sample size formular for descriptive cross-sectional study ^15^ n = (Z^2 pq)/d^2 with the reported proportion of irrational drug use in a teaching hospital of 74.3%,^16^ and provision of 10% non-response. To further improve the validity, this figure was doubled and rounded up to 600 to correspond to the WHO criteria for the prescribing indicator study.^17,18^ For the patient care study, the WHO standard protocol stipulates that at least 100 patient encounters should be recruited for the study.^7^ Therefore, a total of three hundred and thirty (330) samples were selected for the patient care indicator study. The sample of 330 patients was allocated equally to the 10 clinics, while all the forty-eight (48) doctors in the selected consulting clinics were included in the study.

A cluster sampling method with clinical units as sampling frame was used to select ten (10) clinics and their attached pharmacies from the clinical units in the hospital. The selected clinics were Dermatology, Gastroenterology, General surgery, Children Outpatient Clinic (CHOP), Endocrinology, Nephrology, Ophthalmology, Urology, Cardiology and Antenatal clinic. Furthermore, random sampling method was used to select sixty (60) prescriptions from the prescription stumps from each pharmacy attached to these clinics, giving a total of 600 prescriptions. For the patient care indicators study, in line with the WHO guideline of including at least 100 patient encounters in outpatient departments of a single healthcare setting, three hundred and thirty (330) patients (33 patients from each clinic) who met the eligibility criteria and gave their consents were observed in the consulting rooms and pharmacies for the patient care survey. All the forty-eight (48) consulting doctors in the selected clinics who gave their consent were engaged for interview using a semi-structured questionnaire on rational drug use and factors affecting the prescribing practices.

The instruments for this study were the WHO/INRUD drug use forms^17^ or assessing drug use in tertiary hospitals and a semi-structured questionnaire. The data for the prescribing indicators was retrospective. The data was taken from sampled prescription records and recorded manually in structured check list/WHO drug use template accordingly by careful observation. The information extracted include the name of drugs, class of drugs, formulation type, number of drugs per prescriptions, and essential drugs per prescription. Thereafter, the collected data were fed into Statistical Package for Social Sciences (SPSS) version 25 for windows® for analysis. Stopwatches was be used to determine the contact time of health care providers with patient (consultation and dispensing time). Data regarding patient care indicators was prospective. It was taken from patient attendants and their prescriptions in specialist outpatient clinics during the period of data collection and was recorded in the WHO Patient care observational check list/form. Among patient care indicators, data regarding patient knowledge of how to take correct dosage was collected through face-to-face interview and recorded as 1 or 0 for each patient (all or none principle) in accordance with the guideline. The availability of key/essential drugs, EDL was assessed in the pharmacy, and filled in facility indicator form accordingly.

Data to answer the four (4) research questions for this study was generated. Data for Core drug prescription indicators, Patient care indicators, knowledge of patients concerning drug use and dosage and availability of essential drugs in the facilities were analyzed. The Statistical Package for Social Sciences (IBM, SPSS.) was used for analysis of the data. The data was evaluated using the WHO guidelines. Descriptive analyses including frequencies, percentages, means, and standard deviations were conducted and results were presented as texts, illustrated tables and figures.

The WHO’s prescribing indicators were analyzed as follows: ^3^

- Average number of drugs per prescription
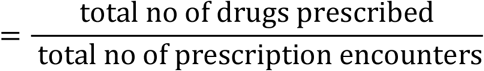
- % of drugs prescribed by generic name
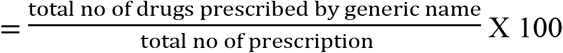
- % of drugs prescribed from EDL
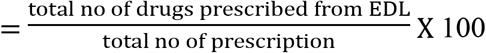
- % of encounters with antibiotics prescribed
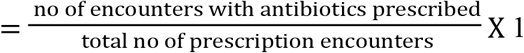
- % of encounters with an injection prescribed
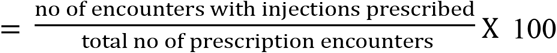

The equations above are used to calculate the core drug indicators according to the WHO standard and the optimal values are shown in Table 1

**Table 1.** Optimal values of core drug indicators

| Core drug indicator | Optimal value |
| --- | --- |
| Prescribing indicators |  |
| Average number of drugs prescribed per patient encounter | 1.6 – 1.8 |
| Percentage of drugs prescribed using generic name | 100 |
| Percentage of encounters with an antibiotic prescribed | 20.0 – 26.8 |
| Percentage of encounters with an injection prescribed | 13.4 – 24.1 |
| Percentage of drugs prescribed from essential drug list | 100 |
| Patient-care indicators |  |
| Average consultation time (minutes) | $\geq 10$ |
| Average dispensing time (minutes) | $\geq 1.5$ |
| Percentage of prescribed drugs actually dispensed | 100 |
| Percentage of dispensed drugs adequately labelled | 100 |
| Percentages of patients with knowledge of correct doses of | 100 |
| Facility-specific indicators |  |
| Availability of essential drug list or formulary to prescribers | 100 |
| Percentage of key drugs available | 100 |

## Results

### Prescribing Indicators

The table 2 above gives the values of the different prescribing indicators. Average number of drugs per encounter was 3.4 ± 1.9 which is twice the WHO standard value of 1.6 – 1.8. The percentages of encounters with antibiotics (40.2%) and injections (24.8%) were higher than the WHO values of 20 – 26.8 and 13.4 – 24.9 respectively. Less than half (43.6%) of the drugs were prescribed by generic names, which is not ideal as the standard is set at 100%, and almost all the prescribed drugs (97.1%) were from the essential drug list.

**Table 2:** WHO/INURD Core Prescribing Indicators of 600 Prescription Encounters of UPTH Pharmacies

Pharmacies
| Prescribing indicator assessed | Value | WHO Standard |
| --- | --- | --- |
| Average no of drugs per encounter | $3.4 \pm 1.9$ | 1.6 – 1.8 |
| Percentage of encounter with antibiotics | 40.2 | 20 – 26.8 |
| Percentage of drugs prescribed by generic | 43.6 | 100 |
| Percentage of encounter with injection | 24.8 | 13.4 – 24.9 |
| Percentage of drugs from EDL | 97.1 | 100 |

### Patient Care Indicators

From the table 3, CHOP clinic has the longest consultation time of 24.40 ± 4.09, while Antenatal clinic (ANC) had the shortest consultation time of 8.40 ± 3.09. Patients from cardiology and gastroenterology clinics received the highest number of drugs prescribed at 3.90 ± 2.07 and 3.90 ± 1.19 respectively, while patients from ANC and general surgery were prescribed the least drugs at 2.20 ± 0.91 and 2.20 ± 1.13 respectively. Patients from gastroenterology also had the highest number of drugs dispensed as 3.90 ± 1.19. The average consulting time of the 10 clinics is 17.46 ± 8.04 minutes, average time to dispense drugs is 7.69 ± 3.80 minutes, average no of drugs prescribed by the consulting doctors after each consultation session is 3.01 ± 1.41 and average number of drugs dispensed by the pharmacies is 2.79 ± 1.30 (92.7%). For drug labelling, 99.8% of the drugs were adequately labelled while most of the patients (82%) understood their drug dosages. The result also shows that there is no copy of the essential drug list available to the doctors in the different clinics and their consulting rooms.

**Table 3:**
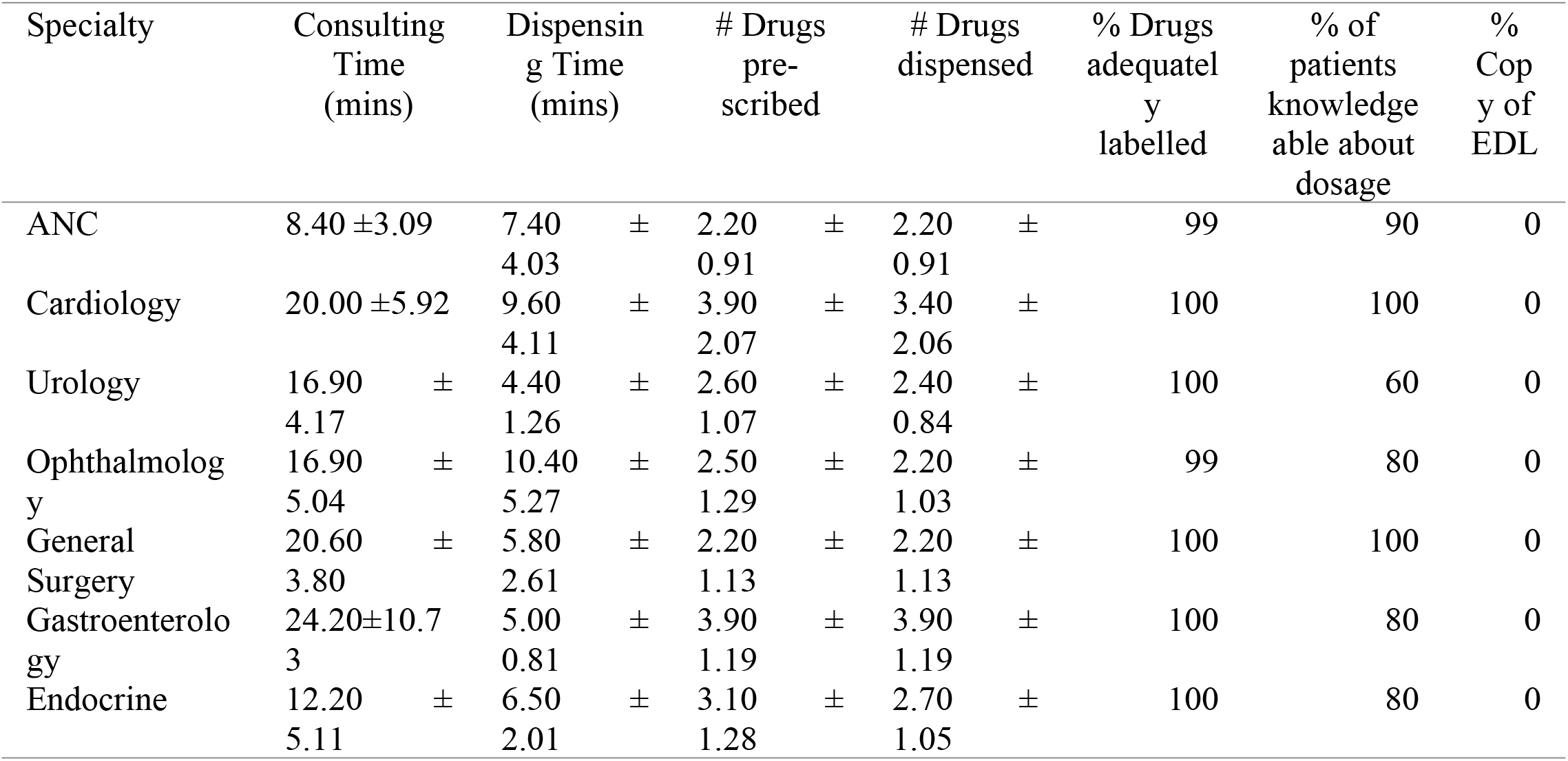
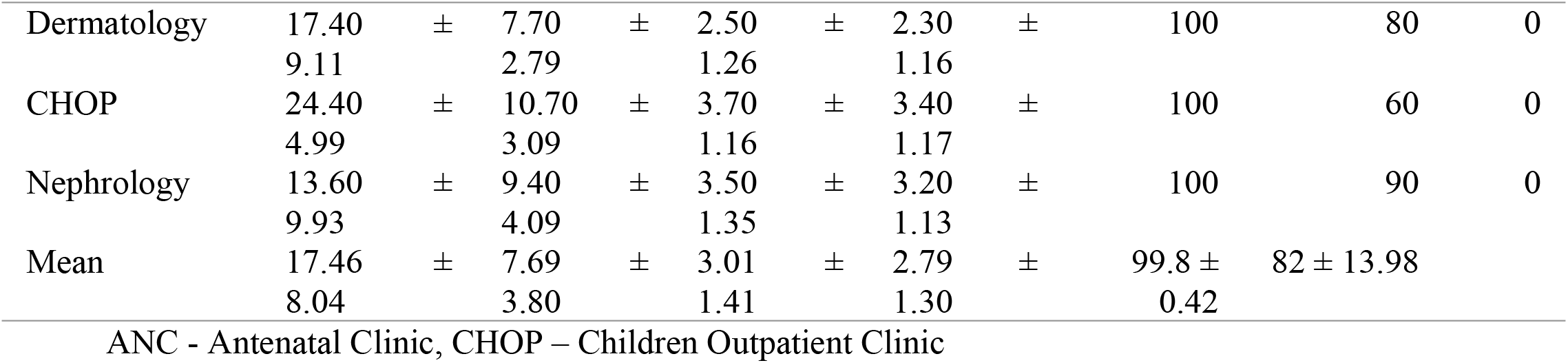
Performance on WHO/INURD Patient Care Indicators in 10 specialty Clinics in UPTH

### Facility Indicator

From table 4, fifteen key drugs according to the WHO were used to assess the availability of key drugs in the facility. The result shows that almost all the key drugs (93.3%) were available and in stock in the UPTH pharmacy.

**Table 4:** Availability of Key Drugs in the pharmacy

| S/N | Key Drug | Availability (Yes/No) | Score (1/0) |
| --- | --- | --- | --- |
| 1 | Oral Rehydration salt | Yes | 1 |
| 2 | Metronidazole/cotrimoxazole | Yes | 1 |
| 3 | Penicillin | Yes | 1 |
| 4 | Paracetamol | Yes | 1 |
| 5 | Artemisinin combination therapy | Yes | 1 |
| 6 | Sulphadoxine pyrimethamine | Yes | 1 |
| 7 | Haematinics | Yes | 1 |
| 8 | Anthelmintics | Yes | 1 |
| 9 | Eye ointments | No | 0 |
| 10 | Skin disinfectants e.g., Gentian violet, iodine etc | Yes | 1 |
| 11 | Skin antifungal agent | Yes | 1 |
| 12 | Non-steroidal anti-inflammatory drugs<br>(NSAIDs) | Yes | 1 |
| 13 | Anti-hypertensive agent | Yes | 1 |
| 14 | Anti-peptic agent | Yes | 1 |
| 15 | Anti-diabetic agents | Yes | 1 |
|  | Total |  | 14 |
|  | Percentage |  | 93.3% |

### Factors affecting prescription practices in UPTH

From the table 5, most respondents agreed to different factors influencing their prescribing patterns. All the respondents (100%) were influenced by the side effects of the drugs, almost all (97.9%) were influenced by the availability of drugs and cost of drugs as key factors affecting their choices of drug prescriptions. No respondent (0%) was affected by patients’ pressure to prescribe drugs and very few (12.5%) prescribed drugs based on patient’s cultural beliefs.

**Table 5:**
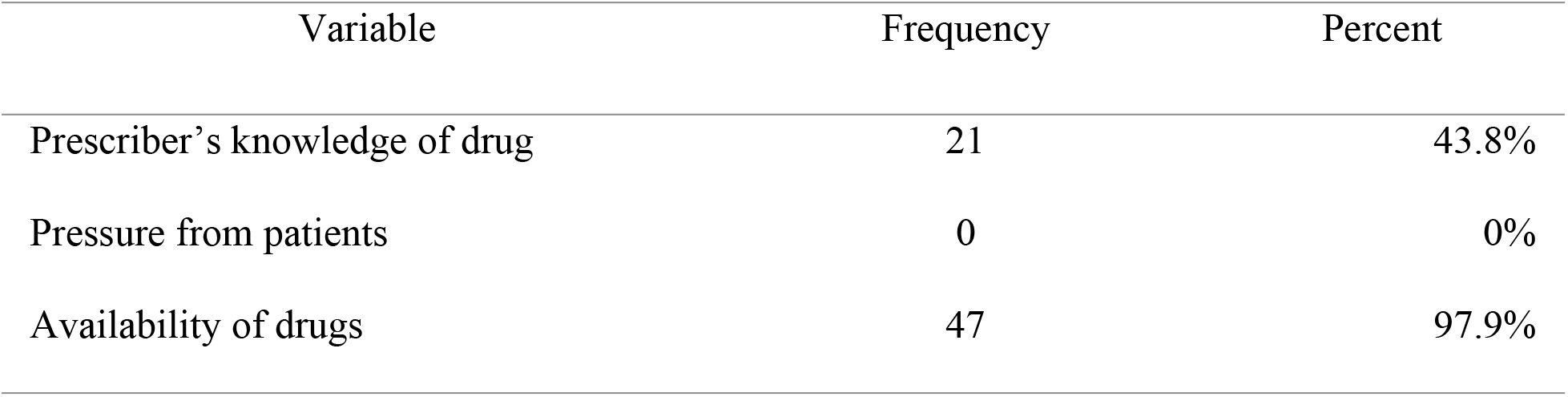
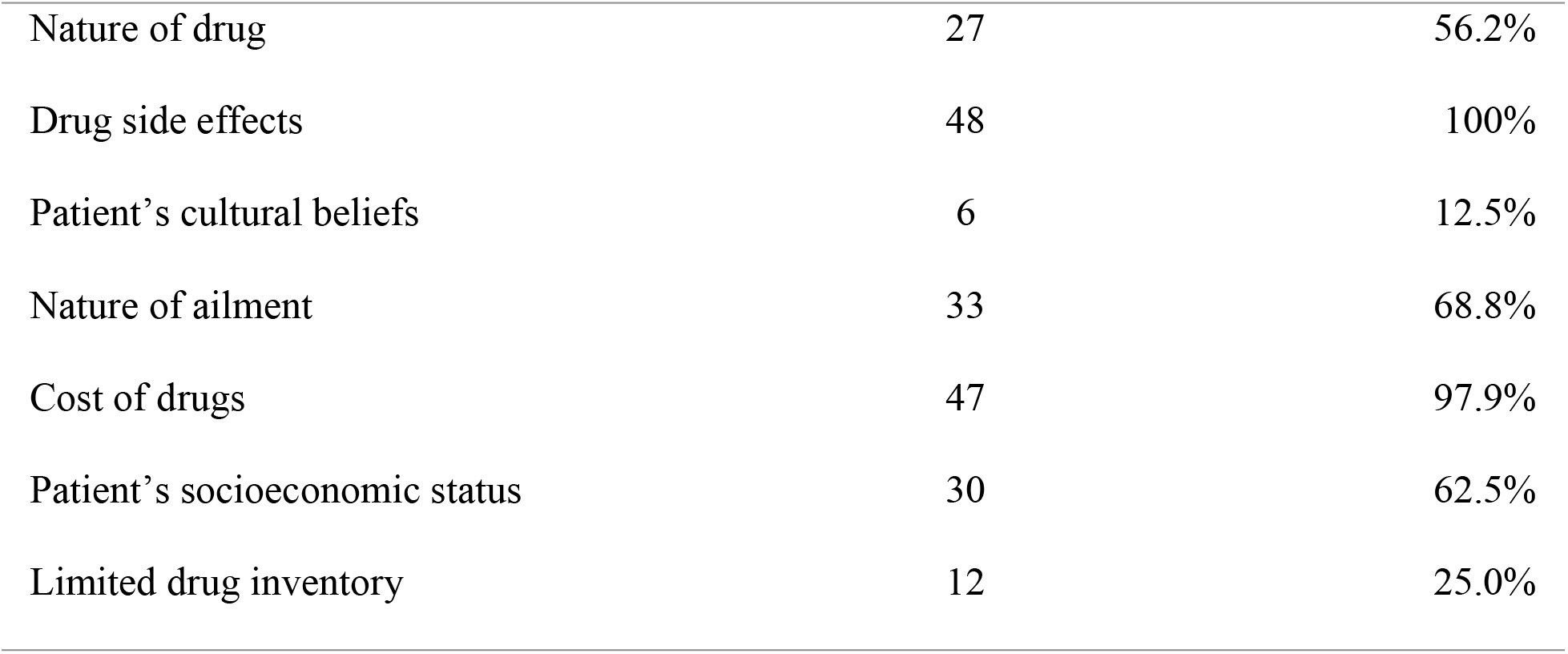
Factors Affecting Prescribing Patterns

### Variable Frequency (48) Percent Knowledge of RDU

From table 6, less than half (43.8%) had a correct knowledge of rational drug use, majority (81.2%) had received no training on rational drug use and majority (66.7%) attributed wrong drug dosage to patients’ mistake. For source of drug information, more respondents (42.9%) agreed to Med/Pharm reps as commonest source of drug information while few (20.8%) agreed to hospital drug bulletin and seminars as source of information.

**Table 6:** Assessment of Rational Drug Use

| Variable | Frequency (48) | Percent |
| --- | --- | --- |
| Knowledge of RDU |  |  |
| Correct | 21 | 43.7% |
| Incorrect | 18 | 37.5% |
| No response | 9 | 18.8% |
| RDU training |  |  |
| Yes | 9 | 18.8% |
| No | 39 | 81.2% |
| Causes of patient's wrong drug dosage |  |  |
| Patient's error | 32 | 66.7% |
| Dispenser's error | 20 | 41.7% |
| Prescriber's error | 10 | 20.8% |
| Don't know | 8 | 16.7% |
| Commonest source of drug information |  |  |
| Hospital drug bulletin | 10 | 20.8% |
| Seminars | 10 | 20.8% |
| Media | 15 | 31.3% |
| Med/Pharm Rep | 23 | 47.9% |
| Internet | 15 | 31.3% |

## Discussion

Findings of this study shows that the average number of drugs per prescription is double of the normal range according to the WHO, with a high rate of antibiotics prescriptions. Some studies in Pakistan^8^ and Central Nepal^10^ also had similar results of polypharmacy and high antibiotics prescribing rates. Similarly, a comprehensive study in the African region by Ofori-Asenso^5^ shows a major deviation of prescribing indicators from the WHO guideline. Several Nigerian studies in different tertiary hospitals also reported cases of polypharmacy, over prescription of antibiotics, low rate of generic prescribing and non-compliance with EDL prescriptions.^19, 20, 21, 22, 23^ The degree of polypharmacy encountered in this study is lower than seen in some studies like Fadare et. al ^24^ whose study showed an average of 8.8 medications per prescription and a very low percentage of generic prescriptions (4.17%). These findings of polypharmacy can adversely affect treatment outcomes predisposing patients to health risks. The increasing rate of antibiotics overuse contributes significantly to the high level of antibiotic resistance in Nigeria and other parts of the world.

Finding showed that doctors in UPTH often practiced branded prescriptions, as less than half of the drugs in the prescriptions were generic prescriptions. Generic prescribing improves understanding and communication between the prescriber and dispenser, thereby leading to effective dispensing.^25^ Also, generic drug prescribing is less expensive than brand prescribing, as the dispenser has the liberty to dispense according to the patient’s level of income. These findings agree with similar studies in the outpatient department of a teaching hospital in Akwa Ibom^23^ which showed low rate of generic prescription, although only 4% of injections were prescribed. The reason for brand prescription recorded in this study stems from the fact that doctors prefer some drug brands and influence of some drug reps makes them prescribe brands from those pharmaceutical companies. Also, cost is another factor that may lead to brand prescription among doctors, as some brands may be cheaper than others.

The EDL was also followed in 97% of the drugs prescribed which is close to the WHO standard of 100%. Also, more injectable drugs were prescribed above the normal WHO range. This agrees to a similar study in GOPD of UPTH which reported an inadequate prescription of injectables and non-compliance with EDL. However, in Jos University Teaching Hospital^21^, injectables were not prescribed routinely (9%) in accordance with the WHO standard with 70% generic prescriptions. Injections have long had a special connotation as particularly powerful and fast acting medicines^26^, this may be a potent reason for the over-prescription of injections in this study. However, Redenna^27^ reported that excessive use of injection predisposes patients to infections.

The findings of this study suggests that the studied specialty clinics in UPTH do not conform to the WHO core indicators as regards patient care. The results of the current study demonstrate that the average consultation and dispensing times were 17.46 min and 7.69 mins, respectively (dispensing time = time when a patient reaches and leaves the pharmacy counter, excluding the waiting time). This result is more than the proposed normal by the WHO. The longer consultation and dispensing times reported in this study can result from the nature of the hospital where training activities takes place with doctors and medical students in the consultation rooms. Also, the consultation process involves physical examination and adequate history taking which improves patient outcome and proper diagnosis. In terms of dispensing time, the hospital process for obtaining drugs from the pharmacy involves costing of the drugs by the pharmacist, payment by the patient and dispensing by the pharmacist. All these takes place around the pharmacy, thereby taking longer time for patients to get drugs. Longer consultation time provides for proper patient evaluation and diagnosis while a long dispensing time provides for good education of the patient on drug related information, which enhances medication adherence.^28^ similar studies in tertiary care facilities in Pakistan^8^ and primary health centres in Brazil, Egypt, Tanzania, Ethiopia, Saudi Arabia and Swaziland all reported short consultation time less than 10 minutes and dispensing time less than 180 seconds.^21^ However, similar studies in tertiary hospitals in Jos^21^ Nigeria, showed an average consultation time of 11.33 minutes and dispensing time of 3.53 minutes which are in accordance with the WHO range and also in line with the findings of this present study.

As regards the labelling of drugs and patients’ knowledge of drug dosage, the result was favorable, as almost all the drugs were adequately labelled, and 82% of the patients had a fair knowledge of their drug dosage. This result is much closer to the required norm of 100%, and higher than other reported cases in different studies in Ethiopia and Pakistan, where 61.8% and 61.6% respectively of patients had adequate knowledge of drug doses.^9^ The studies also reported 86% and 11% respectively of adequately labelled medications. Nigerian studies in Jos showed high degree of patients’ knowledge (93%) and drug labelling (90.4%), while in Kano, 95% of the patients had adequate knowledge with all the dispensed drugs fully labelled. Patient’s knowledge about correct dosage is important to avoid drug overuse and abuse, and further prevent adverse events that affect patients’ health and quality of life. Proper labelling of medications also reduces the incidence of drug misuse by patients as discussed in our literature and previous cited works. This proper labelling and patient’s knowledge reported in this study can be attributed to professionalism and training of the pharmacy dispensers in the hospital pharmacy, who are certified pharmacists.

According to our findings, 92.7% of prescribed drugs were actually dispensed, which is less than the WHO ideal value of 100%. The implication of this shortfall can be attributed to the unavailability of the EDL in some consulting clinics, and unavailability of some key drugs in the pharmacy. This result is higher than similar studies in Jos University Teaching Hospital Nigeria, and Ethiopia^9^, which had 85.3% and 86% respectively of dispensed drugs, while in Pakistan^8^ 97.3% was dispensed out of the total number of prescribed drugs. The unavailability of drugs in the pharmacies reduces the number of drugs actually dispensed to patients out of the prescribed drugs, this forces some patients to seek for drug purchase outside the hospital and in most cases, patronize patent stores where the authenticity of the drugs is not reliable and cases of price hike.

Concerning the facility indicators, 93.3% of the essential drugs were available in the hospital pharmacy, but there was no copy of the essential drug list or hospital drug formulary in the consulting clinics which is not ideal. The reason for this may stem from the fact that the specialist clinics in this study are run by specialist consultants and resident doctors who handle a range of illnesses related to their field and specialty, so they are conversant with the required drugs.

Shortage of any essential drugs in the hospital is disadvantageous for patients in that doctors may not be able to prescribe the correct drugs, or they are limited to prescribing out-of-stock medicines which may pose extra fiscal burden on the patients’ through “out of pocket” expense. Similar studies in Malawi, Ethiopia, Pakistan and Saudi Arabia reported lower values of 67%, 65%, 72.4% and 59.2% respectively for availability of essential drugs in the pharmacies.^8^ Studies based on two Nigerian hospitals reported 90.9% and 62% availability of essential drugs in the two centers respectively^11,12^.

Concerning factors affecting prescribing decision, all the respondents admitted that drug side effects affected their choice of drug, almost all (97%) prescribed based on availability of the drug and drug cost, more than half admitted that the nature of ailment and patients’ socioeconomic status affected their choice of drugs. These findings are critical in rational prescription typified by appropriate indication, drug, patient, information, and monitoring. However, considering the side effects of medications ensures patients safety and optimum health, as every drug have their side effects. The factors considered by UPTH doctors in this study is encouraging and shows that rational drug use is practiced to some extents. Interestingly, no prescriber prescribed drugs based on pressure from patients. This excellent disposition can reduce drug misuse and depicts professionalism on the part of the doctors. These findings are similar to results from two tertiary hospitals in Nigeria which reported drug availability, physician training, drug cost, patient feedback, and patients’ socioeconomic position as major factors influencing prescribing behaviors.^11^ The result seems better than the situation in Tanzanian health centers where doctors’ prescriptions are influenced by excessive patient workload, lack of understanding and patient influence.^12^

Findings from this study shows that less than half of the prescribers understood the concept of rational drug use. This corroborates results obtained from two previous Nigerian studies in Edo and Southeast which reported 23.5% and 25% knowledge respectively.^14^ This poor understanding may be reflective of the level of emphasis on drug use during the mandatory continuing medical education for doctors. Very few prescribers admitted to adequate training on rational drug use which corroborates earlier findings from a teaching hospital in Edo state where the prescribers showed low level of training on rational drug use.

### Implications of the findings

The drug prescribing practices plays a major role in the determination of rational drug use and quality gaps were reported in most of the core drug indicators. These findings have implication for policy, practice, and future research. Firstly, more emphasis should be placed on the regular training of health workers on rational drug use as part of the continuing medical education or professional development. Such training should emphasize enhancing knowledge on the risks and benefits of alternative therapies and adherence to explicit clinical practice guidelines. The hospital should constitute a therapeutic committee with membership drawn from prescribers and dispensers. The team in collaboration with the clinical governance unit (where this exists), should conduct periodic survey to understand the state of drug use in the hospital, to formulate relevant policies for improvement of drug prescription and dispensing practices as well as develop trainings needs on rational drug use for health workers. Essential drug list and treatment guidelines as well as other decision support should be accessible to prescribers to ensure rational and safe use of drugs. There is a preponderance of out-of-pocket payment for receipt of care in the teaching hospital in this setting and drug cost is reported to be a major contributor to the cost of healthcare.^29,30^ The finding that prescribers accord greater consideration to cost than risks and effectiveness of medications during prescription, underscores the need to fast-track the expansion of coverage for social health insurance that would reduce prescription bias, improve rational drug use while not condoning extravagant prescription.

### Study limitation

The study utilized the mutli-dimensional WHO core drug indicators and tools for the conduct of a comprehensive assessment of rational drug use in a typical tertiary hospital setting in Nigeria. However, the cross-sectional design of the study limits causal inferences from being drawn from the findings of this study

## Conclusion

This study evaluated the rational drug use in a Nigerian tertiary hospital setting observed quality gaps in drug use. These results will guide the hospital management in future interventions such as policy enactment, intensification of staff training and strengthening supportive supervision among others to improve the quality of drug prescription and rational drug use in the hospital.

## Data Availability

The data underlying the results presented is attached to this submission

